# A pan-organ exposomic atlas of human aging for precision environmental health

**DOI:** 10.64898/2026.06.26.26356646

**Authors:** Shaopeng Yang, Zhuoyao Xin, Wei Wang

**Affiliations:** State Key Laboratory of Ophthalmology, Zhongshan Ophthalmic Center, Sun Yat-Sen University, Guangdong Provincial Key Laboratory of Ophthalmology and Visual Science, Guangdong Provincial Clinical Research Center for Ocular Diseases, Guangzhou, China; Department of Biomedical Engineering, Johns Hopkins University, Baltimore, Maryland, USA; F.M. Kirby Research Center for Functional Brain Imaging, Kennedy Krieger Institute, Baltimore, Maryland, USA

**Keywords:** exposome, pan-organ aging, aging clocks, age-related disease, health inequity, precision health

## Abstract

Environmental exposures are major modifiable determinants of human aging, yet the evidence remains fragmented across organ-agnostic summaries and rarely confronts population inequity. Here we present an exposomic atlas of pan-organ aging in ∼300,000 UK Biobank adults, mapping 164 environmental and behavioural exposures onto biological aging of the whole body and nine organ subsystems. Comprising 1,476 systematically tested exposure–subsystem associations, the atlas reveals that environmental effects on human aging are pervasively organ-specific, with 65.9% of exposures acting divergently across organ subsystems. This landscape resolves into nine navigable modules that preserve organ selectivity, predict 23 major age-related diseases, and expose distinct dimensions of health inequity. In-silico analyses further show that priorities for ameliorating aging are target-dependent rather than universal, diverge markedly from the whole-body ranking (Kendall’s *τ* = −0.52 to 0.39), reorder substantially across population strata, with findings externally validated in an ethnically distinct cohort. The atlas establishes an organ-resolved and target-aware foundation for precision environmental health.

## Introduction

The environment is the most modifiable determinant of human aging.^1-3^ Decades of epidemiology have catalogued hundreds of physical, behavioural, social, and psychological exposures linked to aging-related phenotypes, yet two gaps continue to limit translation.^3-5^ First, although recent exposome-wide studies have begun to map these influences at scale, the evidence remains fragmented across single-organ and organ-agnostic summaries, leaving unresolved how the exposome organizes across the human organism. Second, the evidence rarely confronts population inequity, even though both exposomic burden and aging susceptibility differ markedly across social and demographic groups. Closing both gaps is a prerequisite for precision environmental health.

Pan-organ resolution is imperative to defining the environmental architecture of human aging. Organs age along divergent trajectories within the same individual, and organ-specific aging captures distinct patterns of disease risk.^6-10^ The same heterogeneity should be expected of how the exposome shapes aging, where it carries an additional structural consequence. Different organs interface with the environment through distinct biological and social pathways, such that a single exposure can act with different magnitudes or even in opposite directions across organs.^5, 11, 12^ Whole-body conclusions can therefore distort the organ-level architecture, making organ heterogeneity a structural constraint of the exposome rather than a description awaiting finer measurement.

Population inequity is the second dimension where averaging fails. Disadvantaged populations are disproportionately exposed to adverse environments, while also weathered by lifelong stress that heightens susceptibility to any subsequent exposure.^13-15^ Yet inequity in causes and inequity in consequences do not necessarily coincide, since the same exposure may differ both in prevalence and in biological impact across populations. Population averages collapse both into a single estimate shaped by dominant groups, producing priorities that systematically underestimate where intervention could yield the largest gain. Environmental priorities calibrated to such averages risk reproducing the very disparities that precision environmental health seeks to address.

Here we present an exposomic atlas of pan-organ aging in ∼300,000 UK Biobank (UKB) adults, mapping 164 environmental and behavioural exposures onto biological aging of the whole body and nine organ subsystems (**Fig. 1**). The atlas shows that system-wide averaging defines an insufficient coordinate that obscures pervasive subsystem heterogeneity. We resolve this landscape into navigable modules that preserve subsystem selectivity and predict major age-related disease risk. We then quantify the dual inequities of exposomic burden and aging consequences across sex, ethnicity, and education, and embed both into the atlas. In-silico analyses derive environmental priorities for aging under specified organ targets and population contexts, with transportability evaluated in an ethnically distinct Chinese cohort.

**Fig. 1.**
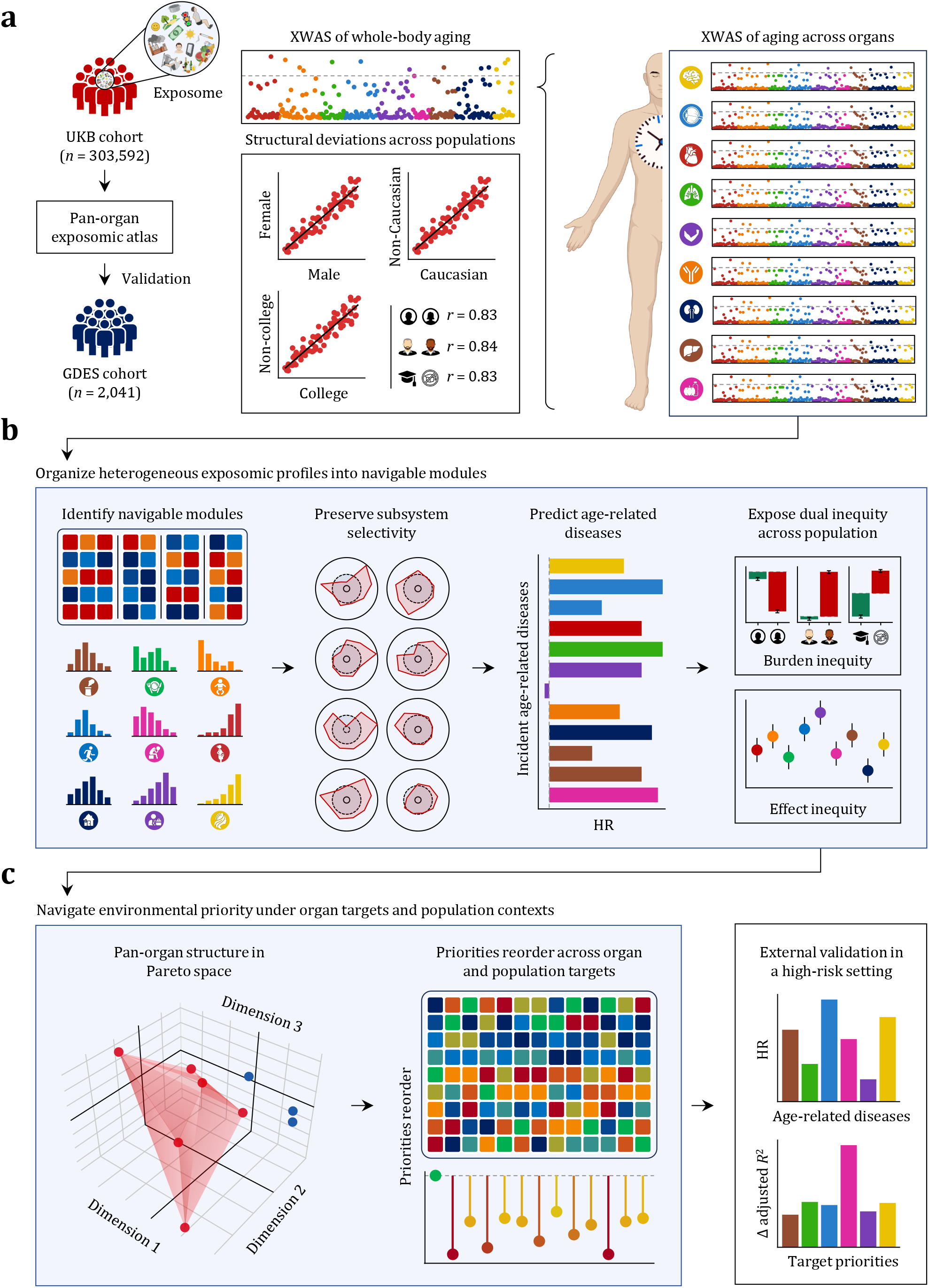
Study overview. (**a**), Discovery and validation design. In the UKB, whole-body aging and organ-specific aging across nine major subsystems were quantified, followed by XWAS of whole-body aging and each organ subsystem. These analyses generated a systematic exposomic atlas of pan-organ aging and established the basis for environmental prioritization. The framework was then evaluated in the GDES. (**b**), Organization of heterogeneous exposomic profiles into navigable modules. Exposures sharing similar pan-organ response profiles were organized into functional modules that compressed the heterogeneous exposomic landscape while preserving subsystem selectivity, anticipating future age-related disease risk, and exposing distinct dimensions of health inequity. (**c**), Derivation of environmental priorities under organ targets and population contexts. Module-level evidence was embedded in Pareto space to represent multisystem trade-offs, allowing priorities to reorder under different target configurations rather than collapse into a single system-wide ranking. Target-specific priorities were then evaluated in an external high-risk cohort (GDES).

## Results

### System-wide aging defines an insufficient coordinate

We first trained machine-learning models in healthy adults free of major medical conditions, using 20-fold cross-validation to estimate biological age for the whole body and each of nine organ subsystems (**Methods**). Consistent with prior work,^6-8^ the gap between estimated and chronological age captures biological aging relative to age-matched peers, yielding normative aging clocks across the organismal hierarchy (**Figs. 2a–b**). The whole-body age gap alone explained 11.1% to 39.8% of the variance across organ-specific age gaps (**Fig. 2b**), indicating a shared trajectory of biological aging captured by system-wide average while leaving substantial subsystem-specific variation unresolved.

**Fig. 2.**
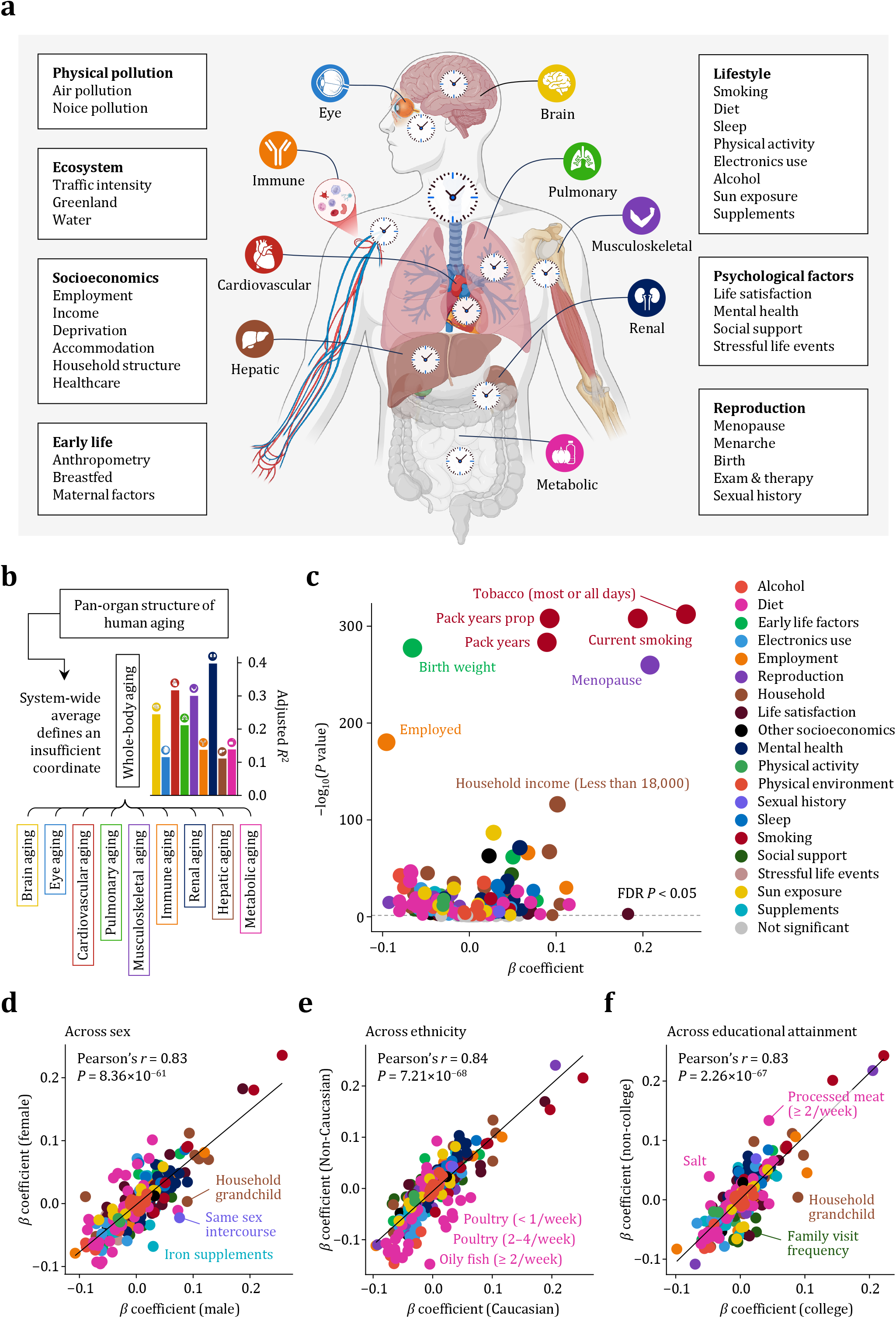
System-wide average provides a necessary but insufficient coordinate for the exposomic architecture of aging. (**a**), Biological aging was estimated for the whole body and nine organ subsystems. Exposome-wide associations were then assessed across seven major exposure domains. (**b**), Pan-organ structure of human aging. Bars indicate the proportion of variance in each organ-specific age gap explained by the whole-body age gap. (**c**), XWAS of whole-body aging (*n* = 283,049). Each point in the volcano plot represents one exposure, positioned by effect size and log-transformed *P* values for the association between that exposure and whole-body aging. Exposures reaching FDR significance are color-coded by category, whereas non-significant associations are shown in grey. The dashed line marks the FDR threshold. The eight most significant exposures are labelled. **(d–f)**, Concordance of exposomic effects on whole-body aging across population strata. Scatter plots compare effect estimates between females (*n* = 188,526) and males (*n* = 152,694) (**d**), Caucasian (*n* = 329,903) and non-Caucasian (*n* = 20,881) participants (**e**), and participants with (*n* = 124,030) and without (*n* = 220,145) a college education (**f**). Each point represents one exposure, and the diagonal line denotes identity. Labeled points mark exposures showing the largest deviations from the shared trend. The *P* values for Pearson’s correlations are shown.

Exposome-wide association study (XWAS) of whole-body aging was performed by serially testing 164 external exposures across seven domains, adjusted for chronological age and major sociodemographic confounders (**Methods**). To reduce reverse-causation bias, exposures indicative of treatment and medication use were excluded. Following multiple testing correction, 128 of 164 exposures (78.0%) were significantly associated with whole-body aging (**Fig. 2c**). Smoking and low income showed the strongest associations with advanced whole-body aging, whereas menopause was the most prominent sex-specific exposure. Higher birth weight and employment were among the strongest correlates with delayed body aging. Additional adjustment for assessment centre had minimal impact on the results (Pearson’s *r*⍰= ⍰0.98).

When stratified by population group, exposomic effects remained broadly concordant across sex (Pearson’s *r*⍰= ⍰0.83), ethnicity (Pearson’s *r*⍰= ⍰0.84), and educational attainment (Pearson’s *r*⍰= ⍰0.83), but retained structured deviations from perfect alignment (**Figs. 2d–f**). Notably, effect modification by educational attainment was comparable in magnitude to that observed across sex and ethnicity, indicating that both biological and social context modulate exposome– aging associations. The largest deviations were observed for same-sex intercourse and iron supplementation across sex (**Fig. 2d**); poultry and oily fish intake across ethnicity (**Fig. 2e**); and processed meat intake and living with grandchildren across education (**Fig. 2f**).

### Atlas of pan-organ aging reveals pervasive heterogeneity across subsystems

Although estimated ages across the nine organ subsystems were correlated, the modest coefficients indicated substantial subsystem heterogeneity (**Fig. 3a**). The pan-organ aging atlas revealed a broad landscape of subsystem-selective exposomic architecture (**Methods**). The number of significant exposures varied markedly across subsystems, ranging from 138 (84.1%) for musculoskeletal aging to 60 (36.6%) for eye aging (**Fig. 3b**). Pack years of smoking and lifespan proportion of pack years were the only exposures with directionally consistent associations across all nine subsystems. By contrast, only 27 exposures (16.5%) showed concordant associations across at least six subsystems, whereas most (65.9%) exhibited clear cross-system heterogeneity.

**Fig. 3.**
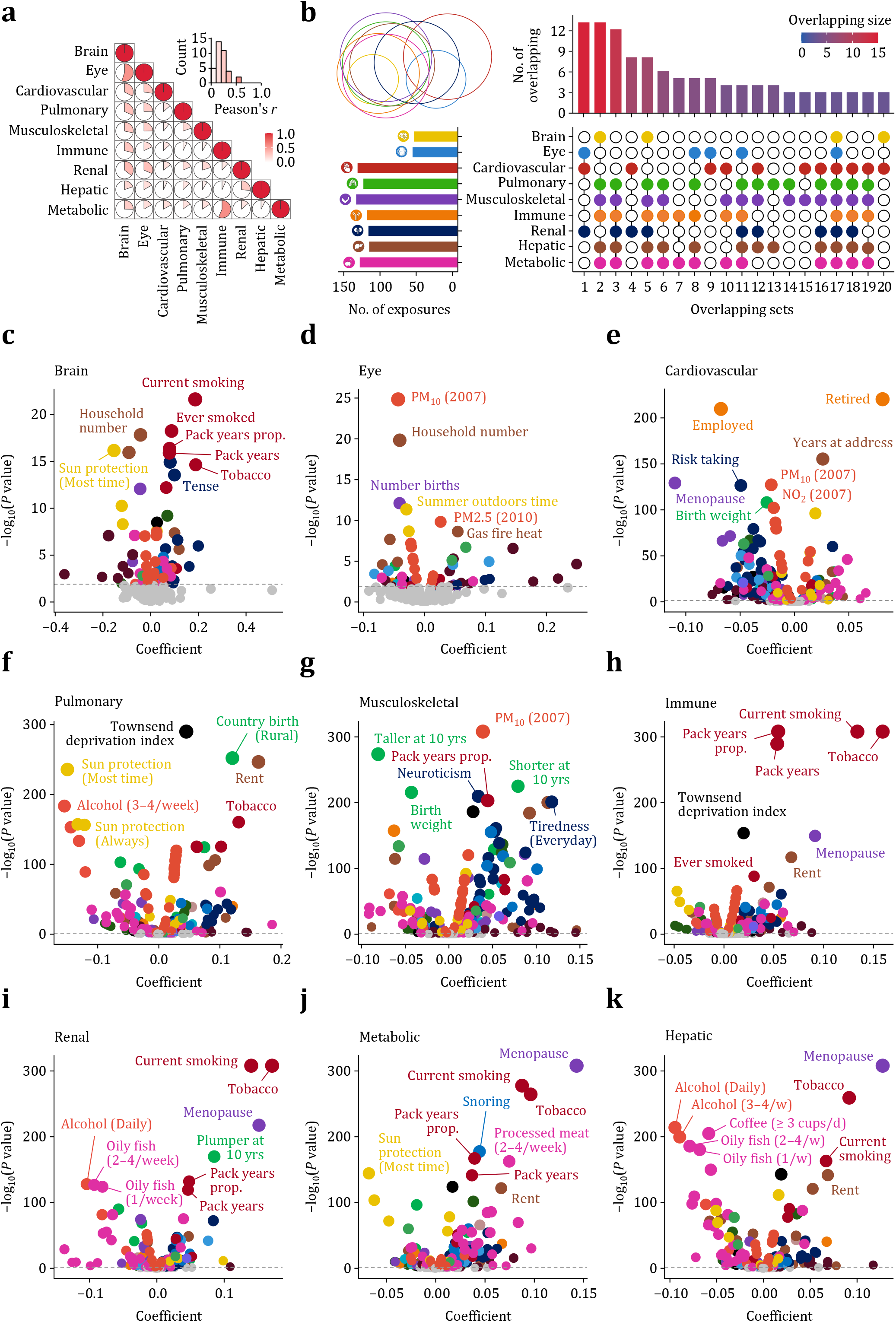
Organ-specific aging reveals pervasive heterogeneity across subsystems. (**a**), Pairwise correlations among estimated age gaps across the nine organ subsystems. Circle size and colour intensity indicate the magnitude of Pearson’s *r*, highlighting modest concordance across subsystems. The inset histogram summarizes the distribution of pairwise correlations. (**b**), Overlap of significant exposome-wide associations across organ subsystems. Left, number of significant exposures identified for each subsystem. Right, the size and composition of overlapping exposure sets across subsystems; connected filled circles indicate the subsystem combination contributing to each overlap set, and bar height indicates the number of exposures in that set. Colour scale denotes overlap size. (**c**), Organ-specific XWAS of pan-organ aging in the brain (*n* = 36,939) (**a**), eye (*n* = 37,322) (**b**), cardiovascular (*n* = 283,049) (**c**), pulmonary (*n* = 283,290) (**d**), musculoskeletal (*n* = 303,592) (**e**), immune (*n* = 298,560) (**f**), renal (*n* = 301,362) (**g**), metabolic (*n* = 297,420) (**h**) and hepatic subsystems (*n* = 284,890) (**i**). Each point represents one exposure, positioned by effect size and log-transformed *P* values for its association with the corresponding subsystem-specific age gap. Exposures reaching FDR significance are color-coded by category, whereas non-significant associations are shown in grey. The dashed line marks the FDR threshold. Top exposures with the strongest subsystem-specific associations are labelled. These results show that although subsystem aging are partially correlated, their exposomic architectures are highly heterogeneous, with both shared and subsystem-selective effects across organs.

Exposures robustly associated with whole-body aging often diverged at the subsystem level (**Figs. 3c–k**). For example, later age at menarche was associated with delayed aging in the whole body and in most organ subsystems, but this association reversed in musculoskeletal aging. This pattern is consistent with prior work linking later menarche to longer lifespan and lower risk across multisystem health outcomes,^16-19^ while supporting a skeletal trade-off whereby delayed pubertal timing reduces cumulative oestrogen exposure and constrain peak bone acquisition.^20, 21^ A similar contrast was observed for childhood height. Greater height was associated with delayed whole-body, cardiovascular, and pulmonary aging, but with accelerated renal aging, consistent with causal evidence linking taller stature to reduced glomerular filtration rate and increased risk of chronic kidney disease.^22-24^

Even when associations were shared across subsystems, their magnitude remained context-dependent (**Figs. 3c–k**). Pack years of smoking and lifespan proportion of pack years were the only exposures associated with accelerated aging across all subsystems, with the strongest effects in brain and pulmonary aging, consistent with the direct interface of inhaled exposure and heightened neurovascular susceptibility. Gym habit showed protective associations across most subsystems, with the strongest effects in musculoskeletal and pulmonary aging, consistent with the role of physical activity in maintaining musculoskeletal and cardiorespiratory fitness.^28, 29^ By contrast, frequent computer gaming was adversely associated with aging across most subsystems, with the strongest effect in the eye, consistent with the direct visual burden of prolonged screen exposure.^30, 31^ We conclude that exposomic effects on pan-organ aging are structured, unevenly distributed, and frequently obscured by system-wide averaging.

### Organizing heterogeneous exposomic profiles into navigable modules

Marked heterogeneity in subsystem-specific exposomic profiles poses a major challenge for translating fragmented associations into actionable priorities. In line with prior work,^4^ some exposures nonetheless shared highly similar pan-organ signatures, but these were not consistently captured by exposomic taxonomy. Smoking, for example, although categorized as a lifestyle exposure, more closely resembled air pollution, whereas sleep-related exposures aligned more closely with psychological than with other lifestyles (**Fig. 4a**). To derive a more decision-relevant structure, we organized exposures into navigable modules by hierarchical clustering of their pan-organ aging profiles (**Methods**). In this profile space, exposures in close proximity shared similar subsystem-specific patterns that resolved into nine coherent modules (**Fig. 4b**).

**Fig. 4.**
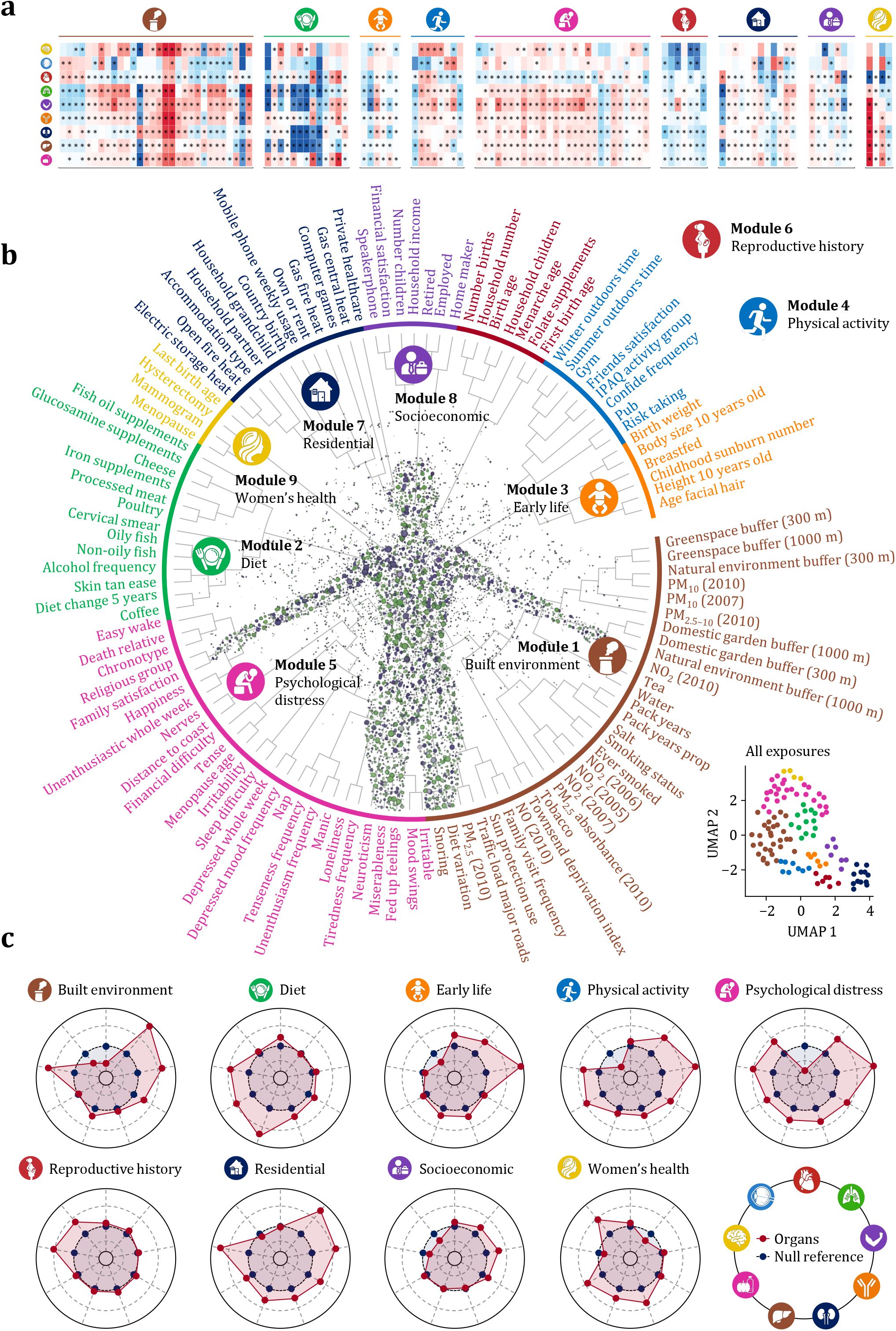
Organizing heterogeneous exposomic profiles into navigable modules. (**a**), Heatmap of subsystem-specific exposomic effect profiles. Rows correspond to the nine organ subsystems and columns to individual exposures. Colour indicates the direction and magnitude of association with subsystem-specific aging, with red denoting positive associations and blue denoting negative associations. (**b**), Hierarchical organization of exposures (*n* = 128) based on pan-organ aging profiles. Coloured labels denote individual exposures, and the outer branches indicate their clustering relationships. The inset shows the two-dimensional UMAP projection of all exposures, illustrating the separation of the nine modules in profile space. (**c**), Module-level pan-organ aging signatures (*n* = 303,592). Radar plots summarize the aggregate subsystem-specific effect profile of each module. Red lines indicate module score associations across the nine organ subsystems, and navy lines indicate the corresponding null reference. These profiles show that the derived modules retain distinct and selective pan-organ aging signatures.

Organized modules preserved pan-organ structure aging while rendering heterogeneous evidence more interpretable at the group level. Module-level associations ranged from five of nine subsystems (55.6%) for socioeconomics to all nine subsystems (100.0%) for psychological distress, yet each module retained a distinct subsystem profile. Built environment was most strongly associated with pulmonary aging, followed by musculoskeletal and brain aging (**Fig. 4c**), consistent with its benefits operating through physical activity and psychological restoration. Other modules were likewise selective, with diet mapping most strongly to hepatic and metabolic aging, early-life factors and physical activity to musculoskeletal aging, and psychological distress to brain aging.

### Organized modules anticipate subsystem-specific age-related diseases

We next tested whether these modules anticipated the incidence of 23 major age-related diseases spanning nine organ subsystems over 16 years of follow-up (**Methods**). Each module was associated with multiple age-related diseases, with 159 significant associations among 207 module–disease pairs (76.8%) after multiple testing correction (**Figs. 5a–i**). Psychological distress showed the broadest disease liability and was associated with increased risk across all 23 diseases examined, followed by built environment, which was associated with 22 of 23 diseases (95.7%). Diet, residential environment, and socioeconomics were each associated with at least one common disease in all nine organ subsystems. These findings remained robust in competing-risk analyses accounting for mortality (**Supplementary Fig. S1**).

**Fig. 5.**
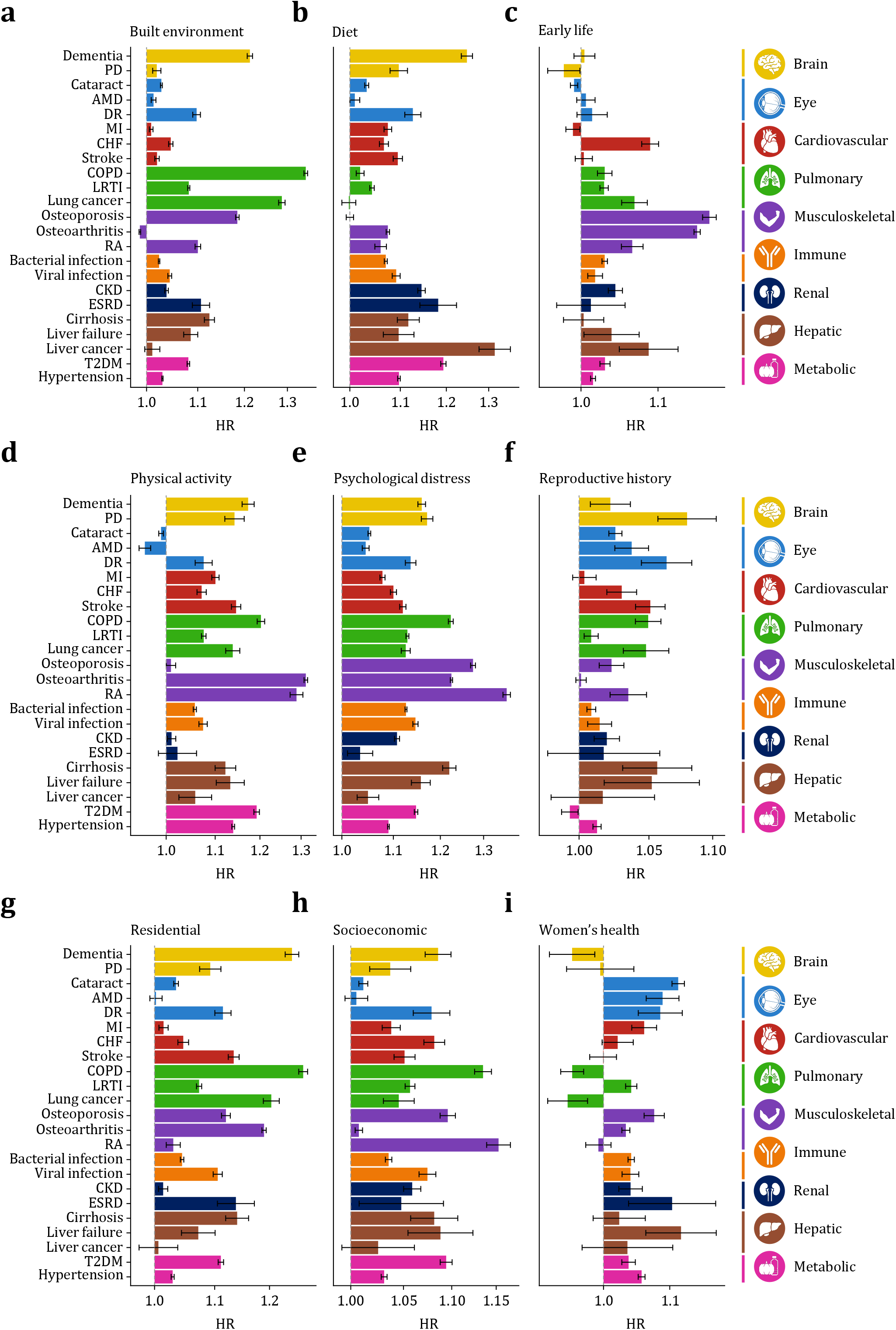
Organized modules anticipate subsystem-specific age-related diseases. (**a–i**), Associations of each exposomic module with incident age-related diseases across nine organ subsystems (*n* = 303,592). Panels show HRs for module–disease associations for built environment (**a**), diet (**b**), early life (**c**), physical activity (**d**), psychological distress (**e**), reproductive history (**f**), residential environment (**g**), socioeconomic (**h**), and women’s health (**i**). Outcomes are grouped and color-coded by subsystem, including brain, eye, cardiovascular, pulmonary, musculoskeletal, immune, renal, hepatic and metabolic diseases. Horizontal bars indicate estimated HRs, and error bars indicate confidence intervals. The dashed vertical line marks the null association. These results show that module–disease associations are non-uniformly distributed across subsystems and are preferentially concentrated in the disease classes emphasized by each module’s selective aging signature.

Module–disease associations were disproportionately concentrated in the subsystems emphasized by each module’s selective aging signature (**Figs. 5a–i**). Built environment was most strongly linked to incident chronic obstructive pulmonary disease (COPD), lung cancer, and dementia, consistent with its dominant associations with pulmonary and brain aging. Diet was most strongly linked to incident liver cancer, dementia, and type 2 diabetes (T2D), matching its selective enrichment in hepatic, brain, and metabolic aging. Osteoporosis, osteoarthritis, and rheumatoid arthritis were among the most prominent outcomes for early-life factors, physical activity, and psychological distress, whereas COPD, dementia, and Parkinson’s disease were also prominent for physical activity and psychological distress.

We conclude that these modules retain subsystem selectivity while linking exposomic architecture to structured patterns of age-related disease risk.

### Sociodemographic inequity shapes both module burden and effects

Because sociodemographic context emerged as a structural modifier of exposomic effects (**Figs. 2d–f**), we next examined dual inequity across sex, ethnicity, and educational attainment. Marked disparities were evident in module burden, most pronounced across ethnicity (**Fig. 6a**). Non-Caucasian groups carried greater burden across all modules except physical activity, for which the difference was not significant. By contrast, burden inequity across sex was module specific: women carried greater burden for adverse built environment, physical activity, psychological distress, residential environment, and socioeconomics, whereas men carried greater burden for adverse diet and early-life factors. Individuals without a college education likewise carried greater burden across most modules.

**Fig. 6.**
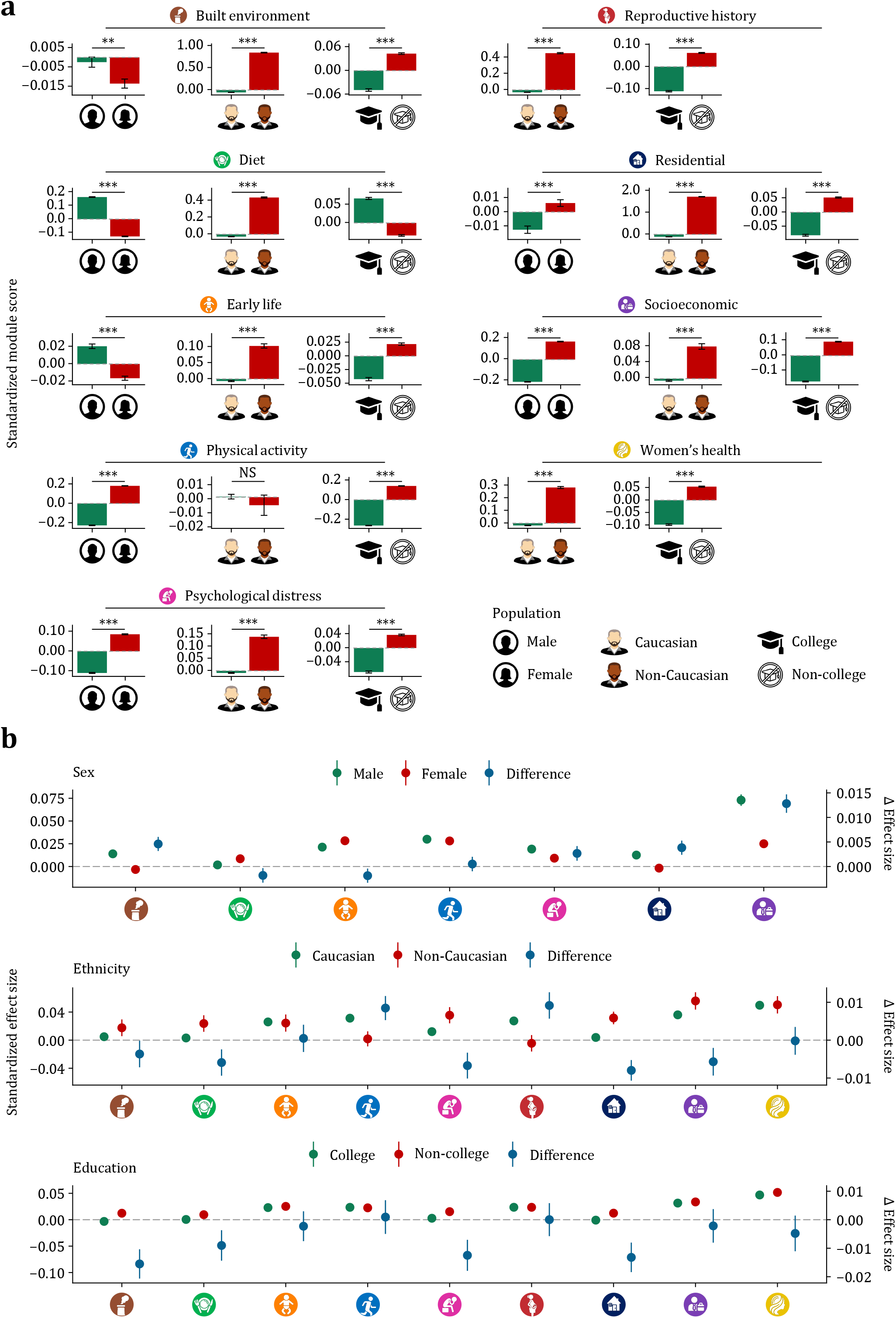
Sociodemographic inequity shapes module burden and module effects. (**a**), Population differences in module burden across sex, ethnicity, and educational attainment. For each exposomic module, bar plots compare standardized module scores between males and females, Caucasian and non-Caucasian participants, and participants with and without a college education. Error bars indicate confidence intervals. The dashed horizontal line denotes no between-group difference. Asterisks denote statistical significance (***P⍰<0.001, **P⍰<0.01, *P⍰<0.05); NS, not significant. (**b**), Population differences in module effect size on pan-organ aging. For each module, points show standardized effect estimates for the two comparison groups across sex, ethnicity, and educational attainment, with blue points indicating the between-group difference. Error bars indicate confidence intervals. The dashed horizontal line denotes no between-group difference. These results show that sociodemographic inequity operates along two distinct dimensions: unequal burden of adverse exposures and unequal aging consequences conditional on burden.

We also observed inequity in the aging consequences of exposomic modules, conditional on their burden (**Fig. 6b**). Across sex, effect inequity remained module specific: although women carried greater burden for adverse built and residential environment, psychological distress, and socioeconomics, men experienced larger aging effects when exposed. By contrast, effect inequity amplified burden inequity across ethnicity and educational attainment. Despite already carrying greater burden across most modules, non-Caucasian groups showed stronger aging effects for adverse built and residential environment, diet, psychological distress, and socioeconomics. A similar pattern was observed across educational attainment, where adverse built and residential environment, diet, and psychological distress exerted stronger effects among those without a college education.

### Priorities reorder under target configurations and health inequity

We next formulated environmental prioritization as a multi-objective optimization problem that integrates subsystem susceptibility, module burden, and population inequity under predefined targets (**Fig. 7a**). Direct comparison of module effects derived from whole-body averaging yielded a single priority ordering that masks subsystem heterogeneity, as reflected by substantial discordance with subsystem-specific solutions (Kendall’s *τ* from −0.33 for brain aging to 0.39 for renal aging; **Fig. 7c**). To resolve this, we mapped exposomic modules into a nine-dimensional Pareto space and identified mutually non-dominant frontiers, where each module represented a conditionally optimal solution under different target configurations (**Methods**). Six modules lay on the Pareto frontier, at least five of which would otherwise have been excluded by any single-objective ranking that enforces a unique solution (**Fig. 7b**).

**Fig. 7.**
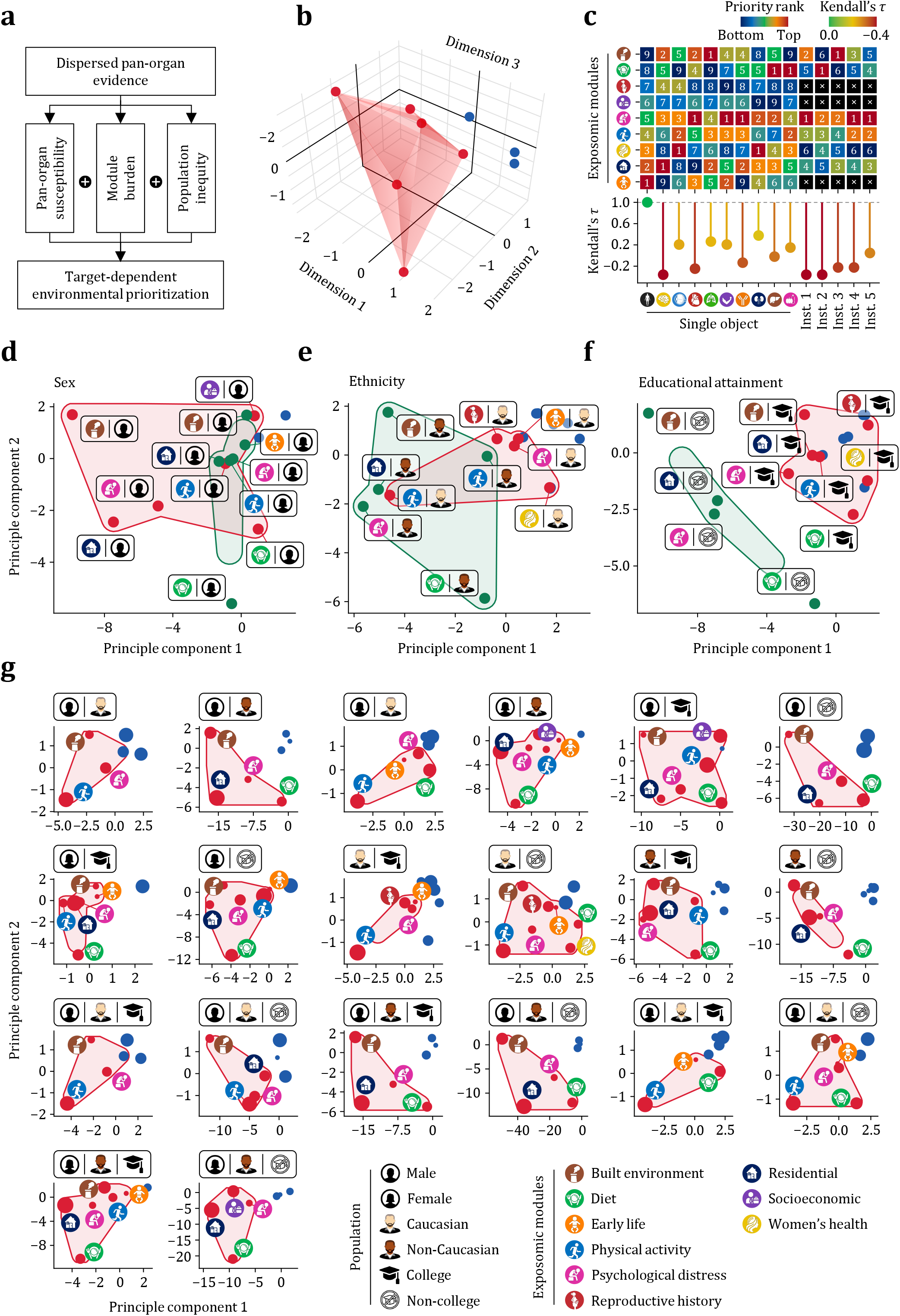
Priorities reorder under target configurations and health inequity. (**a**), Conceptual overview of the priority derivation. Dispersed pan-organ evidence is integrated across three dimensions (subsystem susceptibility, module burden, and population inequity) to derive target-dependent priorities. (**b**), Schematic illustration of Pareto optimization in a reduced decision space. Red points and the shaded surface denote mutually non-dominant solutions on the Pareto frontier, whereas blue points denote dominated solutions. (**c**), Priority rankings of exposomic modules under single-subsystem targets and five representative multisystem target configurations. Heatmap values indicate module rank, from lower priority (blue) to higher priority (red). Crosses denote modules that were Pareto-dominated under multisystem configuration. The lower panel shows Kendall’s *τ* between each target-specific ranking and the ranking derived from whole-body averaging, quantifying the extent of priority reordering. Instance 1, nine-subsystem configuration with equal weights; instance 2, inflammatory–metabolic configuration with equal weights; instance 3, neuro–vascular configuration with equal weights; instance 4, nine-subsystem configuration with musculoskeletal aging weighted twofold; instance 5, inflammatory–metabolic configuration with immune aging weighted twofold. (**d– f**), Projections of population-specific Pareto spaces across sex (**d**), ethnicity (**e**), and educational attainment (**f**). Shaded red and green polygons delineate the Pareto frontiers for the two comparison groups in each analysis, and labelled boxes identify frontier modules within each population group. Blue points outside the polygons denote dominated solutions. (**g**), Projections of Pareto spaces across refined population strata, including pairwise and intersectional combinations of sex, ethnicity and educational attainment. Boxes above each panel indicate the stratum shown. Red polygons delineate the Pareto frontier, with red points marking mutually non-dominant module solutions. Blue points outside the polygons denote dominated solutions. Point size denotes module burden in the corresponding population. Together, these results show that both target specification and population inequity reorder intervention priority.

Different subsystem targets yielded different priority orderings under the same system-wide evidence, reflecting explicit Pareto trade-offs across subsystems (**Fig. 7c**). Equal weighting across all nine subsystems prioritized psychological distress, built environment, and physical activity (Kendall’s *τ* = −0.33). By contrast, an inflammatory–metabolic configuration, reflecting the close coupling of immune, hepatic, and metabolic aging,^32, 33^ elevated diet to the top priority (Kendall’s *τ* = −0.33), whereas a neuro–vascular configuration, motivated by the brain–heart–eye axis,^34-36^ prioritized built environment (Kendall’s *τ* = −0.20). Further upweighting musculoskeletal aging within the nine-subsystem configuration raised physical activity to the second rank, whereas upweighting immune aging within the inflammatory–metabolic configuration shifted psychological distress to the top. By contract, reproductive history, early-life factors, and socioeconomics remained Pareto-dominated.

Population inequity reshaped the Pareto frontier and reordered priority. Because the aging consequences of exposomic modules differed across populations, modules occupied distinct positions in Pareto space. Projections of the high-dimensional frontier revealed clear shifts across sex (**Fig. 7d**), ethnicity (**Fig. 7e**), and educational attainment (**Fig. 7f**), sufficient to alter priority orderings. Under the inflammatory–metabolic configuration, the top priority shifted to physical activity in men (Kendall’s *τ* = −0.14), residential environment in non-Caucasian groups (Kendall’s *τ* = 0.11), and physical activity in those without a college education (Kendall’s *τ* = −0.39). Under the neuro–vascular configuration, the top priority shifted to physical activity in women (Kendall’s *τ* = 0.28), residential environment in non-Caucasian groups (Kendall’s *τ* = 0.17), and built environment in those without a college education (Kendall’s *τ* = −0.22).

Module burden determined actionable impact within Pareto space. Under the neuro–vascular configuration in men, upweighting burden contribution (λ = 2) shifted the top priority from built environment to physical activity (Kendall’s *τ* = 0.14), suggesting that modules with greater burden were preferentially favoured by target-aware prioritization. This effect was amplified across intersecting population strata, where burden and effect inequities jointly reshaped the priority landscape (**Fig. 7g**). Relative to the overall population, the same target shifted priority from psychological distress to physical activity in Caucasian women without a college education (Kendall’s *τ* = −0.24) and to residential environment in non-Caucasian men with a college education (Kendall’s *τ* = 0.05). Even when the top module was unchanged, burden upweighting still reordered priorities, with Kendall’s *τ* values of −0.52 and 0.14 in these two strata, respectively.

To assess whether the identified priorities reflected stable structure rather than stochastic fluctuation, we performed bootstrap resampling under three representative multisystem target configurations (**Methods**). Across resamples, each target reproducibly recovered a distinct priority regime, indicating a highly structured priority landscape (**Table 1**). Leading modules showed high Pareto-membership frequencies and concentrated rank distributions, whereas residual variability was largely confined to neighbouring frontier modules with similar pan-organ profiles that occasionally exchanged nearby ranks. By contrast, modules that were Pareto-dominated, including early-life factors, reproductive history, and socioeconomics, were rarely selected across bootstrap replicates, suggesting that their low priority was structural rather than sampling-dependent.

We conclude that environmental priority is target-dependent rather than universal and is further reshaped by population inequity within a stable pan-organ decision structure.

### External validation supports transportable target-specific priorities

We next asked whether the priority structure derived in the UKB generalized to an independent high-risk population. We therefore evaluated the framework in a prospective cohort of patients with T2D from the Guangzhou Diabetic Eye Study (GDES), followed for six years with baseline pan-organ aging assessment and incident outcome ascertainment (**Methods**). Because only a subset of exposures and organ phenotypes could be harmonized across cohorts, validation was restricted to modules and subsystem targets that could be directly mapped. Pan-organ analysis reproduced the marked cross-subsystem heterogeneity observed in the UKB, with 26 of 28 pairwise Pearson’s r (92.9%) below 0.3 despite shared system-wide aging (**Fig. 8a**).

**Fig. 8.**
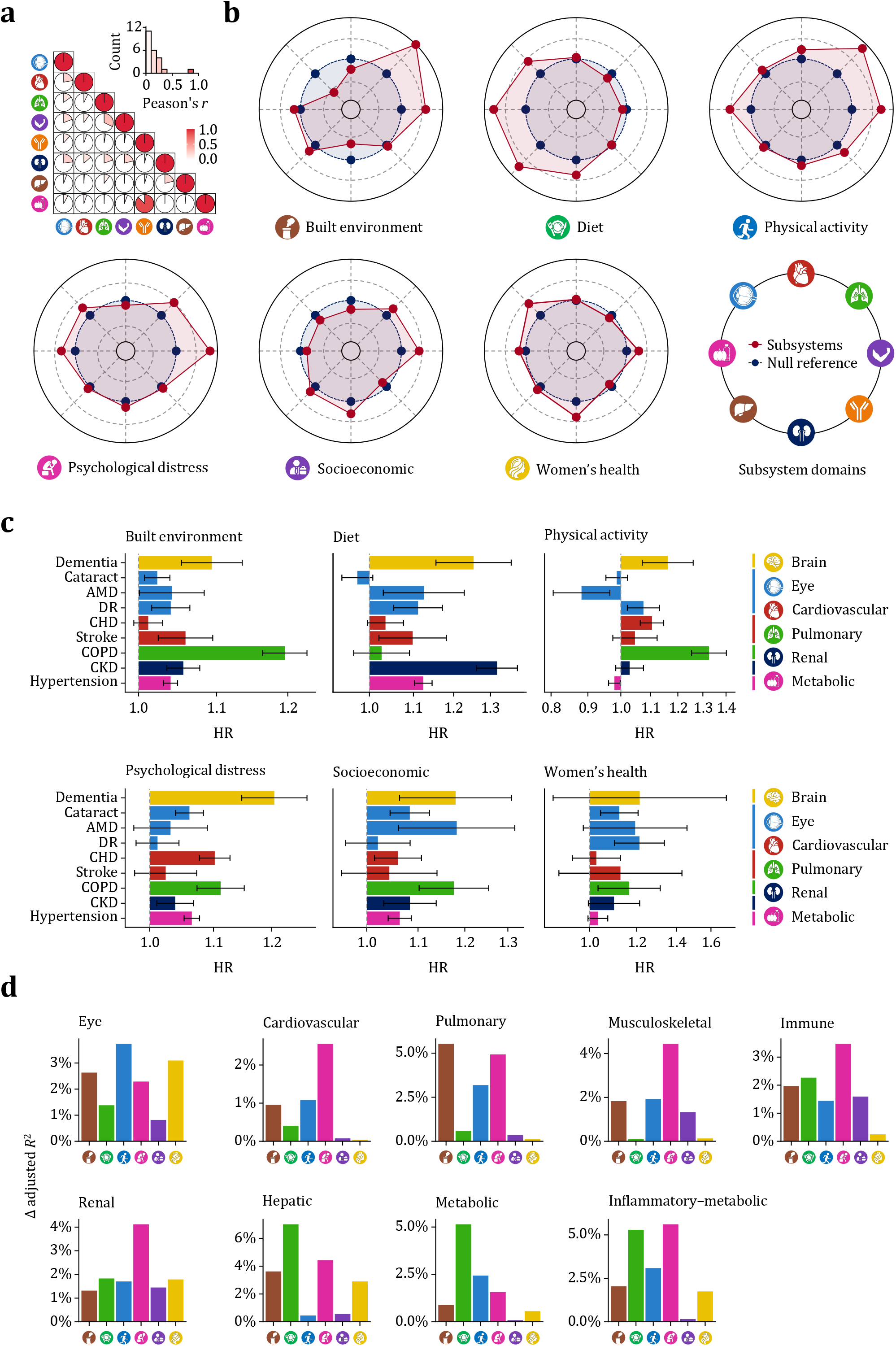
External validation supports transportable target-specific priorities. (**a**), Pairwise correlations among estimated age gaps across the eight organ subsystems harmonized between the UKB and GDES. Circle size and colour intensity indicate the magnitude of Pearson’s r, highlighting modest concordance across subsystems. The inset histogram summarizes the distribution of pairwise correlations. (**b**), Module-level pan-organ aging signatures in the GDES for the six mappable exposomic modules. Radar plots summarize the aggregate subsystem-specific effect profile of each module across the harmonized subsystems. Red lines indicate module score associations, and blue lines indicate the corresponding null reference. (**c**), Associations of mappable exposomic modules with incident age-related diseases in the GDES. Panels show HRs for module– disease associations across brain, eye, cardiovascular, pulmonary, renal, and metabolic outcomes. Horizontal bars indicate estimated HRs, and error bars indicate confidence intervals. The dashed vertical line marks the null association. (**d**), External explanatory performance of UKB-derived module priorities under prespecified subsystem targets. Bars show the additional variance explained in matched subsystem aging in the GDES by each module across individual subsystem targets and the inflammatory–metabolic configuration. The brain aging clock could not be constructed because brain MRI was unavailable. These findings show that subsystem heterogeneity, module selectivity, and target-specific priorities identified in the discovery cohort remain externally informative in a high-risk population.

Projecting modules into the GDES recapitulated the selective aging architecture observed in the UKB. Module effects remained unevenly distributed across subsystems, with built environment and physical activity most strongly associated with pulmonary aging, and physical activity and psychological distress with musculoskeletal aging (**Fig. 8b**). This selectivity extended to clinically relevant outcomes, as modules most strongly associated with specific aging subsystems were also preferentially linked to incident age-related diseases within the corresponding subsystems (**Fig. 8c**). Consistent with the UKB, built environment and physical activity showed the strongest associations with incident COPD, whereas psychological distress was preferentially enriched for incident dementia.

We next tested whether the priority rankings remained externally informative in the GDES. Across single-subsystem targets, the variance in matched subsystem aging explained by each module closely tracked the target-specific priority structure derived from the UKB (**Fig. 8d**). Modules ranked highly in the UKB for a given subsystem generally explained more external variance in the corresponding subsystem, whereas modules prioritized by system-wide average effects were typically not the leading contributors under subsystem-specific targets in the GDES. The same pattern was observed for the inflammatory– metabolic configuration, where explanatory performance was concentrated in diet and psychological distress, consistent with their prioritization in the UKB (**Fig. 8d**).

## Discussion

Here we present an exposomic atlas of pan-organ aging that maps the environmental architecture of human aging across 164 exposures and nine organ subsystems. Three findings define its conceptual contribution. First, organ heterogeneity is the rule rather than the exception, such that whole-body summaries distort the organ-level architecture. Second, this heterogeneity is structured. The atlas resolves into nine navigable modules that retain organ selectivity and anticipate major age-related disease risk. Third, environmental influence on aging is shaped by dual inequities of burden and effect that cannot be recovered from population averages. Together, these findings move the field beyond fragmented description toward an organ-resolved and target-aware foundation for precision environmental health.

Our study responds to a central translational bottleneck in modern epidemiology. As large-scale studies have shifted from testing isolated exposures to mapping exposome-wide landscapes, the evidentiary space has expanded dramatically while the path from association to action has not.^37^ The atlas addresses this gap not by generating more associations, but by giving them an organ-resolved coordinate system in which priorities can be defined, compared, and reordered under explicit targets. Robust association discovery remains essential, yet the field also needs structured representations that make dense evidence navigable. The atlas suggests that the next phase of exposomic epidemiology should advance not only by accumulating evidence, but by building the structures that turn evidence into action.

The pervasive heterogeneity across organ subsystems indicates that environmental intervention cannot admit a universal optimum. since no single strategy can dominate across all organ dimensions.^38, 39^ Within the atlas, optimal priorities are bounded by mutually non-dominant solutions, each gaining priority under a specific organ target while accepting compromise under others. Priorities aimed at musculoskeletal aging may favor physical activity, whereas those aimed at hepatic or metabolic aging may favor diet. Neither is universally superior outside an explicitly specified target. Whole-body averaging is therefore structurally insufficient for prioritization, because it assumes that competing signals can be collapsed onto a commensurable scale without loss of decision-relevant structure. Our results show that this assumption does not hold.

The identified modules function as compressed decision units for the exposome. By resolving shared subsystem-response profiles into modules, they render a high-dimensional exposomic space tractable for interpretation while preserving organ selectivity and remaining clinically informative for major age-related disease risk. Although some within-module components were intuitively coherent whereas others were not, this reflects their role in capturing functional response structure rather than taxonomic similarity. This compression balances tractability with information retention while leaving residual within-module heterogeneity available for hierarchical refinement when finer resolution is required. The framework is therefore scalable across levels of granularity, allowing exposomic evidence to be resolved at different resolutions according to decision context.

Precision environmental health must account for both who is most exposed and who is most affected.^40, 41^ Population-average evidence is shaped by dominant groups and is therefore neither inherently fair nor necessarily transportable. We found that non-Caucasian individuals and those without a college education carried greater adverse exposomic burden across most modules and experienced stronger aging consequences when exposed, implying greater benefit from the same intervention. This pattern is consistent with an equigenic effect, whereby improving health-promoting environments can preferentially reduce health inequality.^42-44^ By quantifying inequity along two distinct dimensions and embedding both into the atlas, our framework allows priorities to be aligned not only to organ targets, but also to the populations most burdened and most affected.

Several limitations should be considered. First, this study captures only a subset of the exposome, as no empirical framework can exhaust the entire exposome. Nor does it capture exposomic dynamics across the life course. Broader and more deeply phenotyped datasets will therefore be needed to extend and refine the framework. Second, we systematically evaluated only linear associations. Future studies that model non-linear exposure–aging relationships may provide greater precision. Third, the discovery cohort was drawn from a Caucasian-skewed population. Although non-Caucasian participants were represented, the limited sample size constrained equally robust subgroup-specific modelling. Nonetheless, external validation in an ethnically distinct cohort supported the transportability of both the module structure and the target-dependent priority beyond the discovery setting.

In summary, the exposomic atlas of pan-organ aging provides an organ-resolved and target-aware foundation for precision environmental health. By moving exposomic inquiry from fragmented description toward structured priority setting, our findings suggest that the next stage of environmental epidemiology will require not only additional associations, but also the architectural structures through which evidence becomes actionable.

## Methods

### Study population

The UKB is a prospective, multicentre cohort study that recruited 502,505 participants from 22 assessment centres across England, Scotland, and Wales between 2006 and 2010.^3, 6, 45^ Participants were 37 to 73 years old at recruitment and underwent extensive questionnaires, physical assessments, and blood and urine assays. The GDES is a community-based cohort study that recruited 2,975 participants with T2D aged 35 to 85 in Guangzhou, China.^46-48^ Baseline assessments were conducted between 2017 and 2019. The study adhered to the Declaration of Helsinki and was approved by the Northwest Multicentre Research Ethics Committee (11/NW/0382) and the Ethics Committee of Zhongshan Ophthalmic Centre (2017KYPJ094). Written informed consent was obtained from all participants before enrolment.

### Age phenotypes and multiorgan aging clocks

Following the phenotype framework defined by Tian et al.,^6^ we selected 78 physical and physiological measures reflecting the function, structure, or general health of the cardiovascular (*n* = 283,049), pulmonary (*n* = 283,290), musculoskeletal (*n* = 303,592), immune (*n* = 298,560), renal (*n* = 301,362), hepatic (*n* = 284,890), and metabolic (*n* = 297,420) systems to construct subsystem-specific aging clocks. In addition, 2,309 structural and functional brain phenotypes and 60 eye phenotypes were used to construct the brain (*n* = 36,939) and eye (*n* = 37,322) aging clocks, respectively.^6, 7^ A detailed data dictionary is provided in **Supplementary Table S1**. All nine subsystem clocks were trained within the UKB using 20-fold cross-validation, following the procedure described by Tian et al.^6^ Missing values were handled using multiple imputation by chained equations with random forests, implemented separately within each cross-validation split.

Specifically, support vector machines (SVM) were trained to predict chronological age from the corresponding phenotypes of each subsystem. In each iteration of the 20-fold cross-validation, a linear SVM was fitted on participants from 19 folds and applied to the held-out fold to generate predicted age for each individual. All predictions were derived exclusively from withheld test sets, thereby eliminating data leakage. All continuous measures were centered and standardized using the mean and standard deviation calculated within the training set before each iteration. For all models, the SVM box constraint and kernel scale were set to unity, and the half-width of the epsilon-insensitive band was set to one-tenth of the standard deviation of chronological age. Models were optimized using sequential minimal optimization with a gap tolerance of 0.001. Age gaps were defined as predicted age minus chronological age and were subsequently residualized by regressing out chronological age.

### Exposures and XWAS

A total of 164 environmental and behavioural variables were included as exposomic exposures in the UKB, following the definition and quality-control framework of Argentieri et al.^3^ These variables comprised baseline exposures that were directly collected or derived at recruitment, had less than 80% missingness, and were available across all assessment centres. All continuous exposure variables were cantered and standardized before analysis, except age at recruitment. Ordinal categorical variables were coded to test linear associations only, without assessing higher-order polynomial contrasts. Nominal categorical exposures were analysed with the most common category as the reference. Multiple-choice questions were recoded as binary dummy variables. A detailed data dictionary is provided in **Supplementary Table S2**.

XWAS of whole-body aging gaps were first performed in the pooled dataset and then stratified by sex, ethnicity, and educational attainment, including male, female, Caucasian, non-Caucasian, college, and non-college groups. To characterize subsystem heterogeneity, XWAS was conducted for each of the nine subsystem-specific aging gaps. In each analysis, associations between individual exposures and aging gaps were assessed serially using linear models adjusted for chronological age, sex, ethnicity, educational attainment, and assessment centre. Sex-specific exposures were analysed within the corresponding sex only. In stratified analyses, the stratifying variable was omitted from the model to avoid singularity. Because UKB participants may underreport alcohol intake as a function of higher disease burden,^49^ self-reported overall health status was additionally adjusted in models for alcohol exposure, following Argentieri et al.^3^ All *P* estimations were two-sided and corrected for multiple testing using the Benjamini-Hochberg method, with adjusted *P*⍰< 0.05 considered statistically significant.

### Navigable modules

To derive a navigable structure of the exposome, we represented each exposure by its pan-organ profile, defined as the vector of effect estimates across subsystem-specific aging gaps. Exposures were grouped by hierarchical clustering of these pan-organ profiles using Euclidean distance, such that exposures within the same module shared similar cross-system patterns. The optimal number of modules was determined by within-cluster sum-of-squares analysis (**Supplementary Fig. S2**), yielding a nine-cluster solution that also showed clear separation on visual inspection of the hierarchical dendrogram. This representation preserved subsystem-selective effect structure while reducing exposomic complexity into interpretable modules. For visualization, exposures were then embedded in a uniform manifold approximation and projection space.

For each module *m*, we next derived a composite score to summarize the participant-level expression of its constituent exposure pattern. Categorical exposures were expanded into dummy indicators, and all exposure variables were standardized prior to aggregation. To assign a coherent polarity within each module, constituent exposures were directionally aligned according to the dominant pan-organ direction of the module itself. Let *b*_*j*_ = (*B*_*j*,1_, … *B*_*j,k*_) denote the pan-organ profile of constituent exposure *j*, where *K* is the number of organ subsystems. For module *m*, we defined its centroid in pan-organ effect space as:

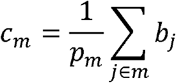

where *p*_*m*_ is the number of constituents in module *m*. Each constituent exposure was then aligned according to its concordance with the module centroid:

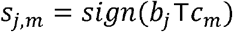

such that exposures pointing in the same pan-organ direction as the module centroid contributed positively, whereas those pointing in the opposite direction contributed negatively. To orient the resulting module score toward greater adverse aging burden, we defined a module-level direction parameter:

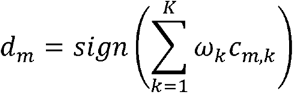

where *w*_*k*_ denotes equal subsystem weights in the primary analysis. For participant *i*, the composite score of module *m*, was calculated as:

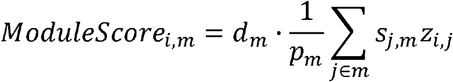

where *z*_*i,j*_ is the standardized value of constituent *j* for participant . Higher module scores therefore indicate greater adverse burden represented by the dominant pan-organ exposure pattern of that module. Associations between module scores and aging outcomes were then assessed using the same covariate adjustment strategy as in the XWAS. All *P* estimations were two-sided and corrected for multiple testing using the Benjamini-Hochberg method.

### Age-related diseases and incident disease analyses

UKB participants were linked to the UK National Health Service (NHS) Hospital Episode Statistics database for hospital admissions and to the NHS Central Registry for mortality. The NHS provides most of the healthcare in the UK, including inpatient and outpatient services, and record linkage was performed using the unique NHS identifier assigned to all residents. Age-related diseases were defined according to the International Classification of Diseases-10, with records available through November 2024. Person-days for each participant were calculated from the date of baseline assessment to the date of disease onset, death, or end of follow-up, whichever occurred first.

Associations between exposomic modules and incident outcomes were assessed serially using Cox proportional hazards models adjusted for the same covariates as in the XWAS. Lifestyle and socioeconomic variables were not included as additional covariates, as they constituted the substantive content of the exposomic modules under evaluation. All *P* estimations were two-sided and corrected for multiple testing using the Benjamini-Hochberg method.

### Multi-objective optimization under organ and population constraints

To translate dispersed pan-organ evidence into structured priorities, we formulated prioritization as a multi-objective optimization problem under explicit subsystem targets and population constraints.^39^ This framework was motivated by the observation that exposomic modules rarely exert aligned effects across organ subsystems. A given module may show a strong effect on one subsystem but weaker, negligible, or even oppositely directed effects on others, such that any single global ranking would collapse intrinsically heterogeneous trade-offs into an artificial unique order. We therefore used Pareto optimization^38, 50, 51^ to preserve the non-dominated structure of the pan-organ effect space.

For each exposomic module *i*, we considered its subsystem-specific effect profile across the *K* organ subsystems. Let *A*_*i,k*_ denote the effect of module *i* on subsystem *k* with positive values indicating more adverse associations with subsystem aging and negative values indicating opposite-direction associations. Module *j* was considered to dominate module *i* if it showed an equal or greater adverse effect in all subsystem dimensions and a strictly greater adverse effect in at least one subsystem *k*^*^:

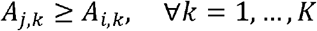

and

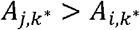

Modules not dominated by any other module were considered Pareto-efficient and together constituted the Pareto frontier. This frontier represents the set of non-dominated candidate modules under multisystem trade-offs, such that no frontier module can be improved in one subsystem without becoming less competitive in at least one other subsystem. By construction, this step preserves modules that may be optimal under different subsystem priorities rather than forcing a single target-agnostic ranking.

To incorporate inequity in exposomic associations across population groups, prioritization was performed within subgroup-specific evidence spaces. Population groups *g* were defined by sex, ethnicity, educational attainment, or their intersections. Population grouping was considered at three levels: (1) one-dimensional grouping, in which sex, ethnicity, and educational attainment were modeled separately; (2) two-dimensional grouping, defined by the pairwise intersections of sex and ethnicity, sex and educational attainment, ethnicity and educational attainment; and (3) three-dimensional grouping, defined by the full intersection of sex, ethnicity, and educational attainment. For each grouping scheme, subgroup-specific module effects were estimated through pooled models with module-by-group interaction terms, thereby allowing the relative positioning of modules in pan-organ effect space to vary across population strata while borrowing strength from the full sample.

For a given population scheme *G* with levels *g*, the effect of module *i* on subsystem *k* within subgroup *g* was estimated from the pooled model:

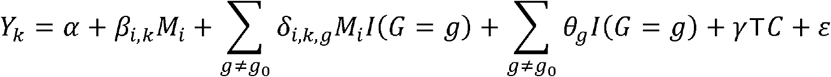

where *Y*_*k*_ is the age gap of subsystem *k. M*_*i*_ is the composite score of module *i*, and *g*_0_ is the reference subgroup, defined as the most prevalent category. Under this formulation, the module effect in the reference subgroup is *β* _*i,k*_ whereas in subgroup *g* the subgroup-specific effect *A*_*i,k,g*_ is *β* _*i,k*_ + *δ*. _*i,k,g*_ Pareto-efficient modules were then reidentified separately within each subgroup-specific effect space. When sex-specific constraints were imposed, modules that were structurally inapplicable to the target population, such as reproductive history and women’s health in male populations, were excluded *a priori* from optimization.

When an explicit decision target was specified, the subgroup-specific Pareto-efficient set was converted into a target-aware ranking. A multisystem target configuration was defined by a subset of subsystem dimensions *T* and a non-negative weight vector *w*, normalized such that ∑_*k* ∈ *T*_ ω_*k*_ = 1. Geometrically, this target specification defines the orientation of a supporting hyperplane in Pareto space, which is translated toward the frontier to identify the preferred solution under the prespecified target. All age gaps and module scores were standardized, such that subsystem-specific effects were expressed on a common scale. Within subgroup *g*, target-oriented susceptibility for module *i* was defined as the weighted aggregation of subgroup-specific module effects across the selected target subsystems:

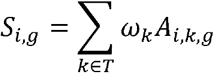

Thus, *S*_*i,g*_ quantifies the net relevance of a module to the prespecified subsystem target under the corresponding population context while retaining the direction of the estimated effects. Equal weights were used by default when no *a priori* target preference was specified, whereas alternative target weights could be introduced to reflect different clinical or public health priorities. The Pareto step and the target-weighting step served distinct roles: the former preserved the set of non-dominated modules under multisystem trade-offs, whereas the latter selected among these candidates according to a specified decision target. Restricting target-aware ranking to the Pareto-efficient set prevents modules that are globally inferior across all subsystem dimensions from being artificially revived by scalar aggregation.

To ensure that modules with non-adverse net effects were not prioritized solely because of high subgroup burden, actionable susceptibility was defined as the non-negative component of the target-oriented susceptibility:

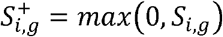

This formulation ensures that a module with large but oppositely directed effects across target subsystems cannot be prioritized unless it shows an overall adverse association with the specified target configuration.

To capture the population-level scale at which a module may be relevant for intervention, we further incorporated subgroup-specific module burden *U*_*i,g*_ into the decision framework. This burden term was defined as the mean adverse-oriented module score within subgroup *g*:

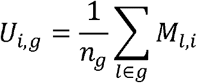

where *n*_*g*_ is the number of participants in subgroup *g*. Because target-oriented susceptibility and module burden are measured on different scales, both components were standardized across modules within each subgroup to 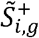 and *Ũ* _*i,g*_ before aggregation. The final priority score was then defined as:

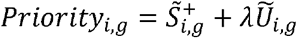

where *λ* ≥ 0 controls the relative weighting assigned to subgroup burden. Under this formulation, *λ* = 1 corresponds to equal weighting of standardized target relevance and standardized subgroup burden on a common scale. In the primary analysis, *λ* was set to 1, and analyses were performed under alternative values to assess the robustness of module rankings to different assumptions regarding the relative emphasis on target susceptibility versus population burden.

To assess whether the identified priorities reflected stable structure rather than sampling variability, we performed bootstrap resampling under three representative target configurations: the nine-subsystem configuration, the inflammatory–metabolic configuration, and the neuro–vascular configuration. In each bootstrap replicate, participants were resampled with replacement from the pooled dataset, the module–subsystem association matrix was re-estimated using the same covariate scheme as in the primary analysis, and exposomic modules were remapped into the corresponding pan-organ effect space. Pareto-efficient modules were then identified, and target-specific priority rankings were calculated using the same target definitions, weighting rules, and burden integration strategy as in the primary framework. This procedure was repeated across 1,000 bootstrap replicates.

For each module under each target configuration, we quantified structural stability using both Pareto-membership frequency and rank-based metrics across bootstrap replicates. We calculated the proportion of replicates in which a module remained Pareto-efficient, together with its median rank, interquartile rank range, and the frequency with which it appeared as the top one or top three priorities. Rank-based summaries were reported only for modules with Pareto-entry frequencies of at least 50% across bootstrap replicates. This restriction was used to avoid optimistic bias in conditional rank summaries, as modules entering the Pareto frontier only sporadically could appear artificially highly ranked when ranks were calculated only among the subset of replicates in which they entered the frontier. Modules with Pareto-entry frequencies below this threshold were therefore interpreted primarily based on frontier-entry stability rather than ordinal rank.

### External validation of target-dependent priorities

Eight of the nine organ-specific aging clocks were reproduced in the GDES; the brain aging clock could not be constructed because brain MRI was unavailable. Six of the nine exposomic modules were also reproduced, whereas early-life factors, reproductive history, and women’s health were unavailable because these exposures were not collected. This resulted in a total of 2,041 available participants. Module scores in the GDES were computed by projecting the harmonized exposure set onto the UKB-derived module structure using the same constituent-level alignment signs and module-level direction parameters as in the discovery cohort. Cross-subsystem correlations, module–subsystem aging associations, and module–incident disease associations were then evaluated using the same analytical framework as in the UKB. Because the GDES is a single-centre cohort with homogeneous ethnicity, adjustment for assessment centre and ethnicity was unnecessary. Across all inter-subsystem, aging, and disease-risk analyses, *P* estimations were two-sided and corrected for multiple testing using the Benjamini-Hochberg method.

For each subsystem target, we quantified the proportion of variance in the corresponding GDES subsystem explained by each module score. Linear models were fitted and the incremental adjusted R^2^ was calculated to represent the additional variance explained after adding module scores to a baseline model including chronological age, sex, duration of diabetes, and HbA1c. Lifestyle and socioeconomic variables were not included in the baseline model, as adjusting for them would partial out the very signals that the validation was designed to assess and would therefore underestimate module-level explanatory performance. These incremental contributions were rank-ordered across modules for each target and compared with the target-specific priority rankings derived from the UKB Pareto analysis. This procedure was applied separately to each single-subsystem target and to the composite inflammatory–metabolic configuration.

## Data availability

All data used in this study are available from UKB via data access procedures (http://www.ukbiobank.ac.uk). Permission to use the UK Biobank Resource was obtained via material transfer agreement as part of Application 105658. The data from the GDES are available upon request to principal investigators Dr. Wei Wang.

## Acknowledgements

This study was funded by the GBRCE for Major Blinding Eye Diseases Prevention and Treatment, the Hainan Province Clinical Medical Center, the National Natural Science Foundation of China (82371086), and the Science and Technology Projects in Guangzhou (SL2024A03J00472; 2025A04J7150). We thank all the participants and staff of the UKB and GDES.

## Author contributions

Study concept and design: W.W.; Acquisition, analyses, or interpretation: S.Y., Z.X., W.W.; Drafting of the manuscript: S.Y., W.W.; Critical revision of the manuscript for important intellectual content: W.W.; Statistical analyses: S.Y., Z.X., W.W.; Obtained funding: W.W.; Administrative, technical, or material support: S.Y., W.W.; Study supervision: W.W.

