## Supplementary material for "A pan-organ exposomic atlas of human aging for precision environmental health": Table 1

Table 1. Bootstrap stability of Pareto membership and target-dependent environmental prioritization across representative target configurations.

| **Exposomic modules** | **Pareto freq. *** | **Median rank (IQR) †** | **Top 1 freq.** | **Top 3 freq.** |
| --- | --- | --- | --- | --- |
| **All subsystems** |  |  |  |  |
| Psychological distress | 100.0% | 1 (1–1) | 99.9% | 100.0% |
| Built environment | 100.0% | 2 (2–3) | 0.0% | 80.7% |
| Physical activity | 100.0% | 3 (2–4) | 0.1% | 70.4% |
| Residential | 100.0% | 4 (3–4) | 0.0% | 48.9% |
| Diet | 100.0% | 5 (5–5) | 0.0% | 0.0% |
| Women’s health | 99.9% | 6 (6–7) | 0.0% | 0.0% |
| Early life | 29.4% | − | − | − |
| Reproductive history | 38.0% | − | − | − |
| Socioeconomic | 14.0% | − | − | − |
| **Inflammatory–metabolic** |  |  |  |  |
| Diet | 100.0% | 1 (1–1) | 99.8% | 100.0% |
| Psychological distress | 100.0% | 2 (2–2) | 0.2% | 100.0% |
| Physical activity | 100.0% | 4 (3–4) | 0.0% | 40.7% |
| Women’s health | 99.9% | 4 (3–5) | 0.0% | 41.3% |
| Residential | 100.0% | 5 (4–5) | 0.0% | 18.0% |
| Built environment | 100.0% | 6 (6–6) | 0.0% | 0.0% |
| Early life | 29.4% | − | − | − |
| Reproductive history | 38.0% | − | − | − |
| Socioeconomic | 14.0% | − | − | − |
| **Neuro–vascular** |  |  |  |  |
| Built environment | 100.0% | 1 (1–1) | 78.8% | 100.0% |
| Psychological distress | 100.0% | 2 (2–2) | 20.4% | 99.3% |
| Residential | 100.0% | 3 (3–4) | 0.7% | 74.7% |
| Physical activity | 100.0% | 4 (4–4) | 0.1% | 24.2% |
| Women’s health | 99.9% | 6 (5–7) | 0.0% | 0.4% |
| Diet | 100.0% | 6 (6–7) | 0.0% | 0.0% |
| Reproductive history | 38.0% | − | − | − |
| Early life | 29.4% | − | − | − |
| Socioeconomic | 14.0% | − | − | − |

* The proportion of 1,000 bootstrap resamples in which a module remained on the Pareto frontier under the specified target configuration. Higher values indicate more stable structural efficiency in multisystem trade-off space.

† Ranks were recomputed after re-estimating the Pareto frontier in each bootstrap resample. Therefore, rank values could exceed the number of Pareto-efficient modules identified in the primary analysis, because boundary modules that were Pareto-dominated in the original sample could enter the frontier in some resamples. Rank-based summaries were reported for modules with Pareto-entry frequencies ≥50%.
