## Supplementary Figures for "A pan-organ exposomic atlas of human aging for precision environmental health"

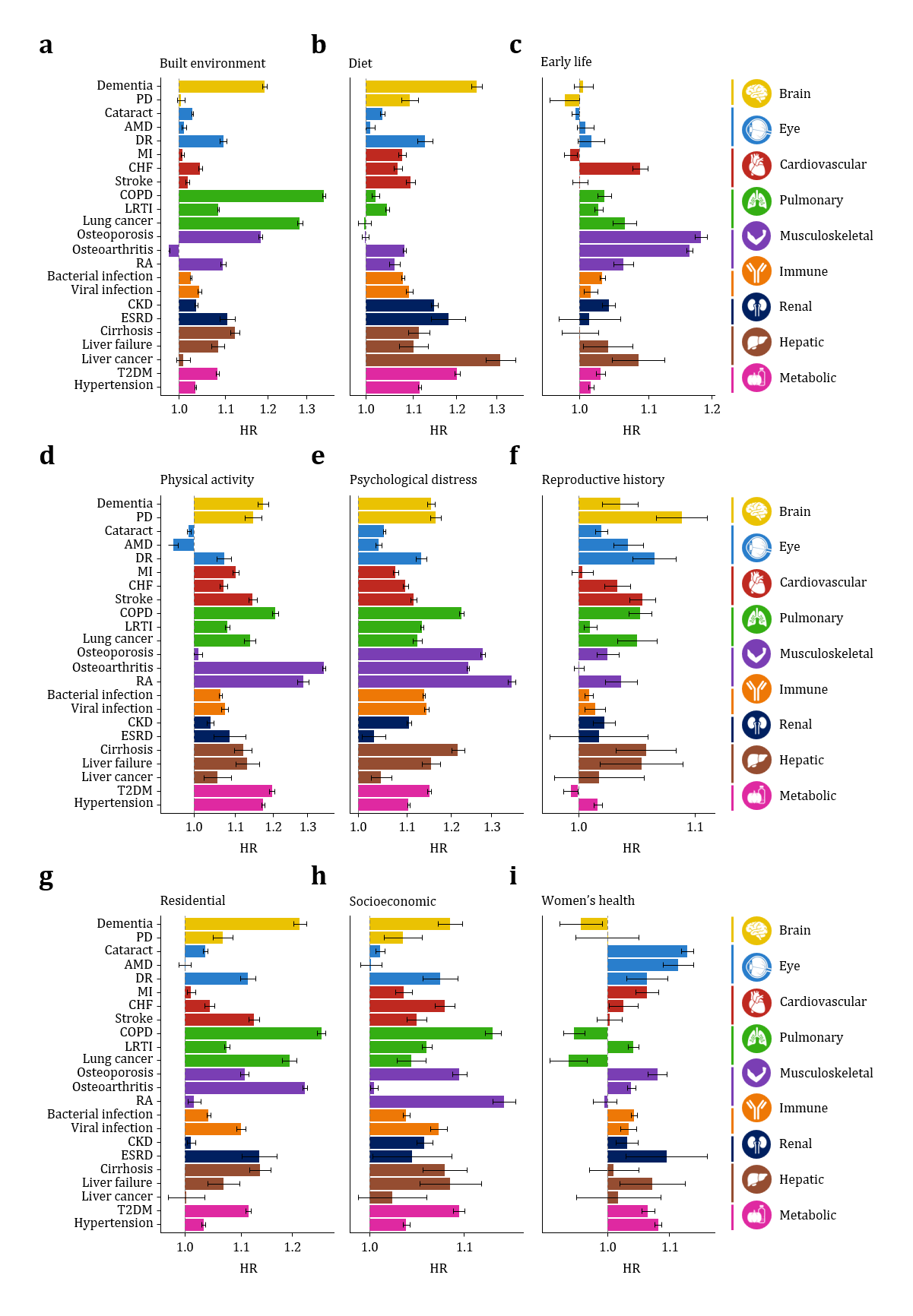

Supplementary Fig. S1. Associations of exposomic modules with risk of incident age-related outcomes **after corrected for competing risk of mortality**.

Findings remained robust after corrected for competing risk of mortality. (**a**), Built environment. (**b**), Diet. (**c**), Early life. (**d**), Physical activity. (**e**) Psychological distress. (**f**) Reproductive history. (**g**), Residential environment. (**h**), Socioeconomics. (**i**), Women’s health.

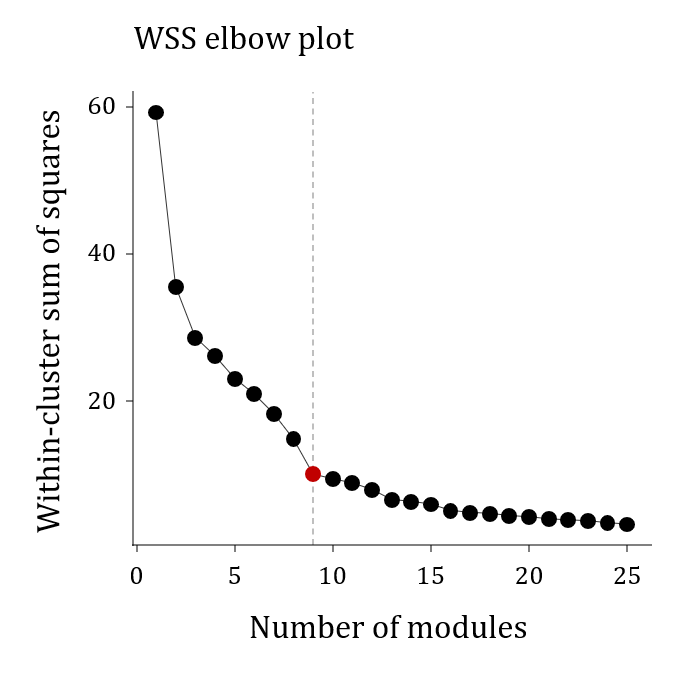

Supplementary Fig. S2. Total within-cluster sum of squares according to the number of **modules**.

An inflection point was identified at nine modules (red dot; dashed line). This choice was further supported by visual inspection of the hierarchical correlation dendrogram, in which the nine-module solution yielded well-defined exposomic groups separated by large branch heights between modules.
